# Clinical, Laboratory, and Imaging Features of 148 Patients with COVID-19 in Bushehr: A Report from the South of Iran

**DOI:** 10.1101/2020.08.11.20172692

**Authors:** Mohsen Keshavarz, Ahmad Tavakoli, Sareh Zanganeh, Mohammad Javad Mousavi, Katayoun Vahdat, Mehdi Mahmudpour, Iraj Nabipour, Amir Hossein Darabi, Saeid Keshmiri

**Author notes:** **Corresponding author** Dr. Mohsen Keshavarz, The Persian Gulf Tropical Medicine Research Center, The Persian Gulf Biomedical Sciences Research Institute, Bushehr University of Medical Sciences, Bushehr, Iran. **Email:**.

## Abstract

**Aim:** To investigate clinical characteristics, laboratory findings, and imaging features of patients confirmed with COVID-19 in Bushehr, a southern province of Iran.

**Method:** During April 29th to May 30th 2020, a total of 148 patients confirmed with COVID-19 infection were admitted to three hospitals in Bushehr province, assigned by the Iranian Ministry of Health.

**Results:** The most common coexisting disease was type 2 diabetes. Levels of ESR, CRP, LDH, and AST among inpatients were higher than the outpatients (*P*<0.05). There were significant differences in the levels of creatinine and BUN between elderly and non-elderly patients (*P*<0.05).

**Conclusion:** Patients with comorbidities and elderly patients are at increased risk of severe progression of COVID-19.

## Introduction

In December 2019, a series of fatal pneumonia cases of unknown etiology was identified in Wuhan City, Hubei Province of China. In January 2020, the Chinese authorities reported that the causative agent was a previously unknown coronavirus, severe acute respiratory syndrome coronavirus 2 (SARS-CoV-2). In February 2020, the World Health Organization (WHO) officially named the disease caused by the SARS-CoV-2 as coronavirus disease-2019 (COVID-19) [1]. Up to July 4, 2020, COVID-19 had been recognized in 216 countries and territories, with a total number of reported laboratory-confirmed cases being 10 922 324 and deaths being 523 011 [2]. Iran is the most affected country by COVID-19 in the Eastern Mediterranean region, and the first cases of the disease was reported on February 20, 2020. The Iranian Ministry of Health (MoH) reported that as of 4 July 2020, there had been 237 878 confirmed cases of the infection and 11 408 deaths in Iran due to COVID-19 [3].

Based on the genome sequence data, it has been revealed that SARS-CoV-2 is a member of the genus Betacoronavirus within the family Coronaviridae [4]. It is an enveloped virus with a single strand, positive-sense RNA genome. Following transmission by inhalation of respiratory droplets, contact with contaminated surfaces, or fecal-oral route, and other means, viral replication takes place in cells of the respiratory and gastrointestinal tracts. The average incubation period of the disease is reported to be 2-14 days, however, there is evidence that it can last as long as 19-27 days [5].

The clinical manifestations of COVID-19 can range from asymptomatic mild disease to severe respiratory failure requiring hospital admission and mechanical ventilation [6]. The main clinical symptoms in COVID-19 patients are fever, cough, fatigue, and sore throat [7]. Although, many studies have been published regarding the clinical characteristics of COVID-19 patients from different countries, there are only few published from Iran. Therefore, this study aimed to investigate clinical, laboratory, and imaging findings of patients infected with COVID-19 in Bushehr, a southern province of Iran.

## Patients & methods

### Study design and population

During April 29th to May 30th 2020, a total of 900 patients suspected with COVID-19 infection were admitted to the Shohadaye-Khalije-Fars Hospital, Nabi Akram Health Center, and Shahid Ganji. These three designated hospitals are responsible for the management of COVID-19 patients in Bushehr province, Iran, assigned by the Iranian Ministry of Health. Patients were divided into two groups, inpatients and outpatient. Both oropharyngeal and nasopharyngeal specimens were collected for each patient and combined into a viral transport medium (VTM) to be tested as a single sample for SARS-CoV-2. Samples were transferred to the virology department of Bushehr University of Medical Sciences under refrigeration. The study was approved by Ethics Committee of Bushehr University of Medical Sciences (ethic code number: IR.BPUMS.REC.1399.026), and written informed consent was obtained from each participant before enrollment.

### Data Collection

Patients were diagnosed with COVID-19 infection, according to the criterion of the WHO interim guidance [8]. For patients with confirmed COVID-19 infection, blood tests were performed, and all data including laboratory results, epidemiological and demographic characteristics, clinical signs and symptoms, and underlying conditions were obtained from the patients. Routine blood examinations were erythrocyte sedimentation rate (ESR), complete blood count (CBC), serum biochemical tests (including blood urea nitrogen, Creatinine, Creatine Phosphokinase, lactate dehydrogenase, aspartate aminotransferase, alanine transaminase, C-reactive protein). Computed tomography (CT) information and patients’ outcomes (death or discharge) were also evaluated. CT findings were classified as ground glass opacities (GGOs), consolidation, consolidation with GGOs, pleural effusion, ground glass and pleural effusion and normal by an expert radiologist.

### Statistical analysis

Statistical analysis was performed using Statistical Package for the Social Sciences (SPSS) for Windows release 25.0 (SPSS Inc., Chicago, IL, USA). Descriptive statistics (mean, SD, CI) were generated for demographic and clinical characteristics. Normal distribution of parameters was analyzed using Kolmogorov-Smirnov test. Since the distribution of the variables did not follow a normal distribution, non-parametric tests were applied. Mann-Whitney U test was used to compare the measurements between groups. Odds ratio (OR) with a 95% confidence interval (CI) was used to evaluate the effect and association of qualitative data. P-values <0.05 were considered to indicate significant statistically.

## Results

### Demographic characteristics

During April 29th to May 30th 2020, a total of nine hundred cases suspected with COVID-19 infection were admitted to our hospitals. Among them, one hundred forty-eight patients (16.4%) were confirmed to have COVID-19 infection by real-time reverse transcription polymerase chain reaction (rRT-PCR); 53 (35.8%) of those 148 patients were outpatients and 95 (64.2%) were inpatients. The ages of patients with COVID-19 infection ranged from 1 to 90 years (mean age of 44.49 ± 17.4 years), and over half of the patients (52.7%) were men. The mean age of the subjects in the inpatient and outpatient groups were 48.89±17.69 and 38.00±15.78, respectively (*P*<0.001). According to the sex distribution, inpatients were composed of 48 (50.5%) males and 47 (49.5%) females. However, 30 (56.6%) males and 23 (43.4%) females were included in the outpatient group (**Table 1**).

**Table 1.**
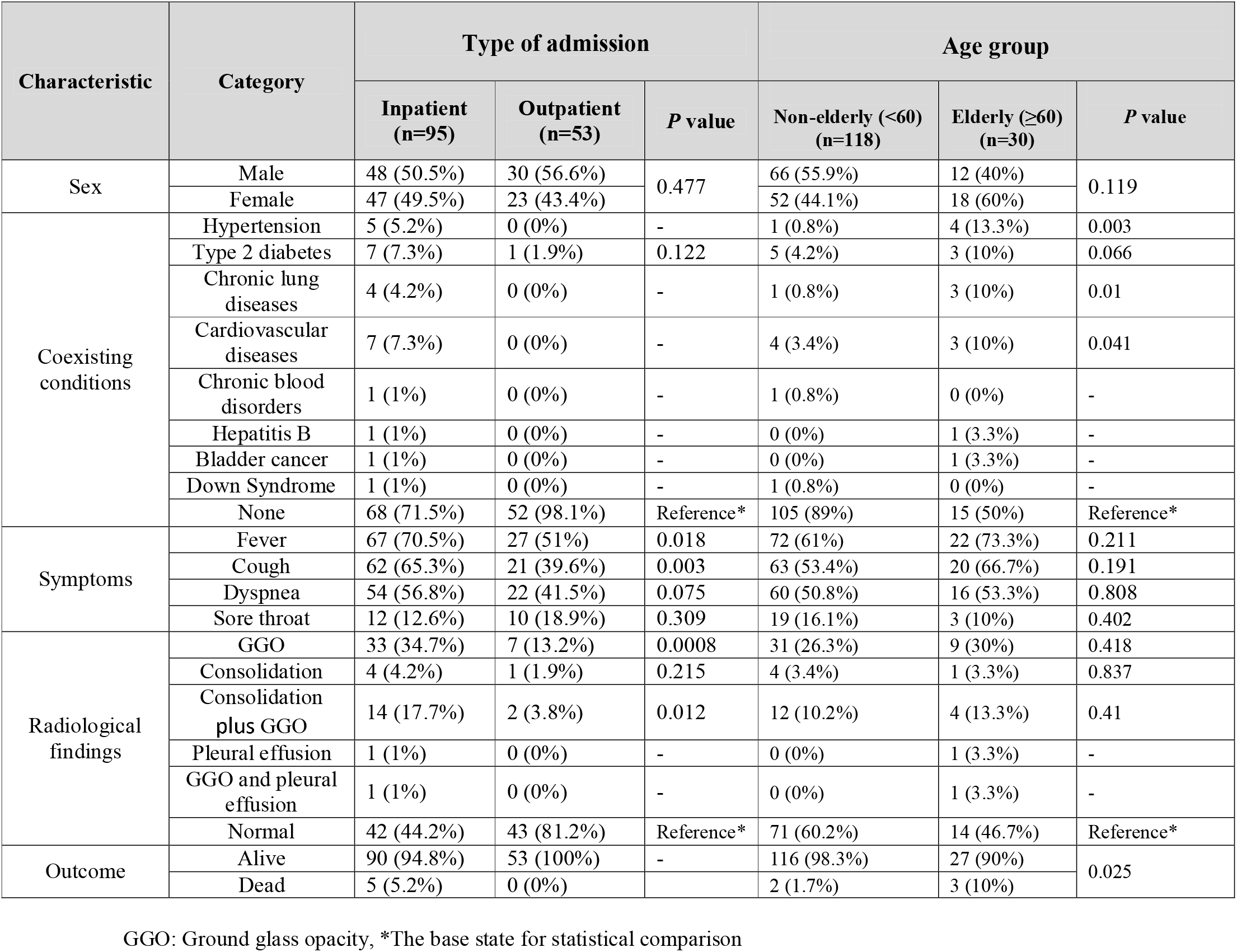
Association of the baseline data and clinical manifestations of the study subjects based on the type of admission (inpatient or outpatient) and age group

### Comorbidities and clinical manifestations

Inpatients showed underlying medical conditions more frequently than the outpatients. The most common coexisting medical conditions in the inpatient group were type 2 diabetes (n=7; 7.3%) and cardiovascular diseases (n=7; 7.3%), followed by hypertension (n=5; 5.2%) and chronic lung diseases (n=4; 4.2%). Among outpatients, only 1 (1.9%) patient had type 2 diabetes, whereas 52 patients (98.1%) did not present underlying diseases (**Table 1**).

Fever was the most common clinical presentation among both inpatients and outpatients (70.5% and 51%, respectively). The cough was the second most frequently observed symptom among inpatients, found in 65.3% patients, while it was observed in 39.6% of outpatients. Dyspnea was more common among inpatients (56.8%) than in outpatients (41.5%) (**Table 1**).

### Laboratory and radiographic findings

Considering the blood leukocytes, the count of WBCs had no significant difference between two groups (6.60±3.31 *vs*. 6.41±2.12, *P*= 0.39). However, lymphocyte counts were lower in the inpatient group in comparison to the outpatient group (1.36±0.76 *vs*. 2.04±0.90, *P*< 0.0001). With respect to the inflammatory markers, ESR was significantly higher in the inpatients in comparison to the outpatients (44.99±26.48 *vs*. 17.95±17.97; *P*< 0.0001). Additionally, CRP level was significantly higher among the inpatients than the outpatients (49.02±35.72 *vs*. 15.49±24.55; *P*< 0.0001; **Table 2**). Both BUN and creatinine levels were higher in the inpatients in comparison to the outpatients, but the differences were not statistically significant (*P*= 0.84 and *P*= 0.48, respectively). Moreover, CPK levels did not have significant change between two groups (*P*= 0.41). However, LDH level was higher among the inpatients than the outpatients (559.57±380.54 *vs*. 358.97±119.83, *P*< 0.0001). Among the liver enzymes, AST level was significantly higher in the inpatient group compared to the outpatient group (38.84±38.77 *vs*. 26.16±13.66; *P*< 0.001). However, no statistically significant difference was detected between two groups (36.77±45.99 *vs*. 30.16±22.38, *P*= 0.39; **Table 2**).

**Table 2.** Baseline data and paraclinical findings of the study participants.

| Characteristic | Type of admission |  |  | Age |  |  |
| --- | --- | --- | --- | --- | --- | --- |
|  | Inpatient (n=95) | Outpatient (n=53) | *P* value* | Non-elderly (n=118) | Elderly (n=30) | *P* value* |
| **Age** (year) | 48.89±17.69 | 38.00±15.78 | <0.001 | 38.03±12.6 | 70.15±8.14 | <0.0001 |
| **WBC** (10<sup>9</sup>/L) | 6.60±3.31 | 6.41±2.12 | 0.39 | 6.42±2.84 | 6.94±3.3 | 0.882 |
| **Lymphocyte** (10<sup>9</sup>/L) | 1.36±0.76 | 2.04±0.90 | <0.0001 | 1.69±0.92 | 1.27±0.58 | 0.002 |
| **ESR** (mm/h) | 44.99±26.48 | 17.95±17.97 | <0.0001 | 29.52±23.98 | 56.65±27.25 | <0.0001 |
| **CRP** (mg/L) | 49.02±35.72 | 15.49±24.55 | <0.0001 | 31.62±34.22 | 57.09±35.39 | <0.001 |
| **BUN** (mg/dL) | 17.36±13.26 | 14.09±4.28 | 0.84 | 15.12±10.96 | 20.1±10.69 | <0.001 |

|  |  |  |  |  |  |  |
| --- | --- | --- | --- | --- | --- | --- |
| **Creatinine** (mg/dL) | 1.13±0.67 | 0.98±0.23 | 0.48 | 1.01±0.45 | 1.3±0.82 | 0.003 |
| **CPK** (IU/L) | 192.98±244.41 | 138.35±148.30 | 0.41 | 175.62±229.36 | 166.84±165.08 | 0.958 |
| **LDH** (U/L) | 559.57±380.54 | 358.97±119.83 | <0.0001 | 480.01±351.58 | 520.11±226.51 | 0.049 |
| **AST** (U/L) | 38.84±38.77 | 26.16±13.66 | <0.001 | 35.45±36.25 | 30.5±13.93 | 0.969 |
| **ALT** (U/L) | 36.77±45.99 | 30.16±22.38 | 0.39 | 36.67±43.4 | 26.46±16.83 | 0.146 |

WBC; white blood cell, ESR: erythrocyte sedimentation rate, CRP; C-reactive protein, BUN; blood urine nitrogen, CPK; creatine kinase, LDH; lactate dehydrogenase, AST; aspartate aminotransferase, ALT; alanine Aminotransferase
\*Non-parametric, Mann–Whitney U was used for between groups analysis.

Regarding age group, we observed a significant increase in the levels of ESR among the elderly patients (56.65±27.25 mm/h) compared to the non-elderly patients (29.52±23.98 mm/h) (*P*<0.0001). Also, the levels of CRP were significantly different between these groups, so that a higher level was observed among the elderly patients (57.09±35.39 mg/L) than the non-elderly ones (31.62±34.22 mg/L). Similarly, higher levels of BUN and LDH were seen in the elderly patients compared to the non-elderly patients (**Table 2**).

According to the chest CT examinations, ground glass opacities were observed in 33 (34.7%) inpatients and 7 (13.2%) outpatients, and the difference was statistically significant (*P*= 0.0008). Additionally, consolidation in conjunction with ground glass opacities were detected in 14 (17.7%) inpatients and 2 (3.8%) outpatients, and the difference was statistically significant (*P*= 0.012). In the inpatient group, 5 (5.2%) death cases were seen, while no death case was detected in the outpatients (**Table 1, Figure 1**).

**Figure 1.**
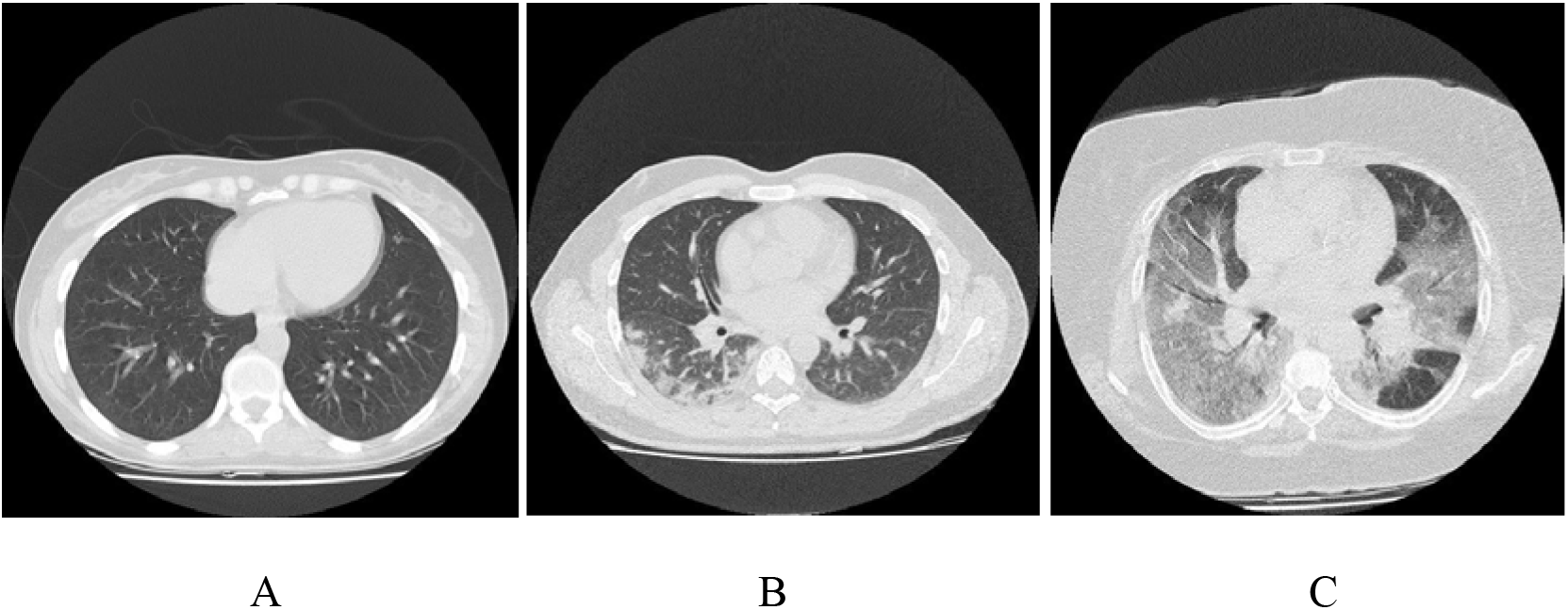
Representative CT Images of patients with COVID-19. A: Female, 27 years old (Outpatient) with fever and dyspnea. Axial CT image showed normal lung markings; B: Female, 35 years old (inpatient with alive outcome) with fever and dyspnea. Axial CT image showed small bilateral areas of groundglass opacities; C: Female, 60 years old (inpatient with death outcome) with cough and dyspnea, Axial CT image shows larger ground-glass opacities in the bilateral.

## Discussion

In the face of the great threat posed by COVID-19 to human health, laboratory assessment and early prognosis of the patient’s condition should be given more attention. Currently, the rapid spread of COVID-19 in Bushehr, a province in southern Iran, has led to the dramatic increase in the number of hospitalized patients as well as increased mortality in the recent weeks. In this descriptive study, we evaluated 148 patients confirmed with COVID-19. Patients were divided into the outpatient and inpatient groups, and their clinical symptoms, laboratory parameters, and chest CT features were evaluated.

Since SARS-CoV-2 is a novel pathogen, the pre-existing immunity is not present in the human community, which makes all humans susceptible to the infection. In high-risk groups such as elderly people and individuals with co-existing comorbidity, outcome of infection with SARS-CoV-2 will be more serious [9]. Our results showed that among underlying medical comorbidities, type 2 diabetes, cardiovascular diseases, and hypertension were the most common in all patients with COVID-19. In addition, frequency of comorbidities was higher in the inpatients compared to the outpatients. These results are in consistent with the findings of another study conducted by Nikpouraghdam et al. in Iran [10]. They found that diabetes, hypertension, and cardiovascular diseases are amongst the most common comorbidities in COVID-19 patients. In another investigation performed by Norooznezhad et al, on 431 patients with a definite diagnosis for COVID-19 infection, underlying medical comorbidities such as cardiovascular diseases and diabetes were the most frequently identified comorbidities [11]. Shahriarirad et al. carried out a study on 113 hospitalized confirmed cases of COVID-19 admitted to hospitals in Shiraz [12]. Similar to our findings, hypertension, diabetes, and cardiovascular diseases were the most prevalent underlying comorbidities among hospitalized patients with COVID-19.

Laboratory parameters are pivotal in helping us to identify COVID-19 cases, and can use in understanding of the disease course, prognosis, and the decision-making process. As a prominent result, we observed a significant decrease in lymphocyte counts among the inpatients compared to the outpatients, and the decrease was more remarkable in elderly patients. In parallel with our findings, several recent studies have been suggested that lymphopenia is a strong indicator of infection with COVID-19 [13-16]. According to the results of a recent meta-analysis, COVID-19 patients presenting lymphopenia have an approximately 5-fold increased risk to develop severe form of the disease [17]. Several potential mechanisms leading to the lymphocyte depletion have been proposed such as direct infection and death of lymphocytes by the virus, destroying lymphatic organs, and lymphocytes inhibition by metabolic molecules produced by metabolic disorders [18].

Levels of ESR, CRP, LDH, and AST among inpatients were higher than the outpatients in our study, and the results are in accordance with the previous studies. It has been documented that elevated LDH levels are associated with worse outcomes in COVID-19 patients [19]. One of the striking findings of the present study is that there was no significant difference in the levels of creatinine and BUN between the inpatients and outpatients, while the difference was observed between elderly and non-elderly patients. This finding indicates that COVID-19 infection is associated with more severe renal failure and abnormalities in elderly patients than non-elderly ones.

Our findings showed that the prevalence of GGO and pulmonary consolidations were significantly higher among inpatients compared to outpatients. This is in line with the results of the recent meta-analysis conducted by Sun et al. They reported that GGO, consolidation, and GGO plus consolidation were the most common findings reported in 94.5% of the studies [20]. There are several limitations to our study that should be considered. First, due to the study design, some important laboratory parameters and biomarkers were not measured in our patients, such as D-dimer, B-type natriuretic peptide (BNP), N-terminal pro b-type natriuretic peptide (NT-proBNP), and Procalcitonin. Second, because of the small sample size in the study, our analyses are low in power. Third, number of patients under the age of 18 were limited, therefore, we could not perform any analysis on pediatric patients with COVID-19.

## Conclusions

In summary, this study reported the clinical, laboratory, and imaging characteristics of 148 patients confirmed with COVID-19 infection in Bushehr, a southern province of Iran. The mean age of patients in the inpatient group was significantly higher than the outpatients. Also, inpatients had underlying diseases more frequently than the outpatients. The most common underlying diseases in the inpatient group were type 2 diabetes, cardiovascular diseases, hypertension and chronic lung diseases. Fever and cough were the most common symptom observed among inpatients. Ground glass opacities were observed in 34.7% of inpatients and 13.2% of outpatients. Furthermore, consolidation plus ground glass opacities were detected in 17.7% of inpatients and 3.8% of outpatients. In patients with typical manifestations and imaging changes, lymphopenia can be used as a potential indicator for diagnosis of COVID-19. Moreover, analysis of BUN and creatinine levels in serum of elderly patients showed that COVID-19 is associated with impaired renal function and related abnormalities.

#### Summary points

- During April 29th to May 30th 2020, a total of 148 patients confirmed with COVID-19 infection were admitted to three hospitals in Bushehr, a province in the south of Iran.
- Among the patients, 35.8% were outpatients and 64.2% were inpatients.
- The mean age of inpatients and outpatients was 48.89±17.69 and 38.00±15.78, respectively.
- The frequency of underlying disease among inpatients was higher than outpatients.
- The most common underlying disease in all patients was type 2 diabetes.
- Ground glass opacities were observed in lung CT images of 34.7% of inpatients and 13.2% of outpatients.
- Levels of ESR, CRP, LDH, and AST among inpatients were higher than outpatients.
- A significant increase was seen in the levels of ESR, BUN, CRP, and LDH among elderly patients compared to non-elderly patients.

## Data Availability

All data generated or analyzed during this study are included in this article

## Acknowledgements

This work was financially supported by a grant (Grant No. 1572) from the Bushehr University of Medical Sciences.

## References

1. Song P, Li W, Xie J, Hou Y, You C. Cytokine Storm Induced by SARS-CoV-2. Clin. Chim. Acta. 509, 280–287 (2020).

2. World Health Organization (WHO). Coronavirus disease 2019 (COVID-19). Situation Report – 166. Internet. Available from: https://www.who.int/docs/defaultsource/coronaviruse/situation-reports/20200704-covid-19-sitrep-166.pdf?sfvrsn=6247972_2.

3. COVID-19 kills another 148 Iranians over past 24 hours. Available from: https://en.irna.ir/news/83843849/COVID-19-kills-another-148-Iranians-over-past-24-hours. (July 4, 2020).

4. Gu H, Chu DK, Peiris M, Poon LL. Multivariate analyses of codon usage of SARS-CoV-2 and other betacoronaviruses. Virus Evol. 6(1), 1–10 (2020).

5. Sarvepalli D. Coronavirus Disease 2019: A Comprehensive Review of Etiology, Pathogenesis, Diagnosis, and Ongoing Clinical Trials. Cureus. 12(5), 1–10 (2020)

6. Li Y, Shi J, Xia J et al. Asymptomatic and Symptomatic Patients With Non-severe Coronavirus Disease (COVID-19) Have Similar Clinical Features and Virological Courses: A Retrospective Single Center Study. Front. Microbiol. 11, 1-8 (2020).

7. Singhal T. A review of coronavirus disease-2019 (COVID-19). Indian J. Pediatr. 87(4), 281–286 (2020).

8. World Health Organization (WHO). Laboratory testing for coronavirus disease 2019 (COVID-19) in suspected human cases: interim guidance, 2 March 2020. World Health Organization; 2020.

9. Guan W-j, Liang W-h, Zhao Y et al. Comorbidity and its impact on 1590 patients with Covid-19 in China: A Nationwide Analysis. Eur. Respir. J. 55(5), 1–14 (2020)

10. Nikpouraghdam M, Farahani AJ, Alishiri G et al. Epidemiological characteristics of coronavirus disease 2019 (COVID-19) patients in IRAN: A single center study. J. Clin. Virol. 127, 1-4 (2020)

11. Norooznezhad AH, Najafi F, Riahi P, Moradinazar M, Shakiba E, Mostafaei S. Primary Symptoms, Comorbidities, and Outcomes of 431 Hospitalized Patients with Confirmative RT-PCR Results for COVID-19. Am J Trop Med Hyg. Online ahead of print (2020).

12. Shahriarirad R, Khodamoradi Z, Erfani A et al. Epidemiological and clinical features of 2019 novel coronavirus diseases (COVID-19) in the South of Iran. BMC Infect. Dis. 20(1), 1-12 (2020).

13. Cheng Z, Lu Y, Cao Q et al. Clinical features and chest CT manifestations of coronavirus disease 2019 (COVID-19) in a single-center study in Shanghai, China. AJR Am. J. Roentgenol. 215(1), 121–126 (2020).

14. Li Y, Wang W, Lei Y et al. Comparison of the clinical characteristics between RNA positive and negative patients clinically diagnosed with 2019 novel coronavirus pneumonia. Zhonghua Jie He He Hu Xi Za Zhi. 43(5), 427–430 (2020).

15. Guan W-j, Ni Z-y, Hu Y et al. Clinical characteristics of coronavirus disease 2019 in China. N. Engl. J. Med. 382(18), 1708–1720 (2020).

16. Rodriguez-Morales AJ, Cardona-Ospina JA, Gutiérrez-Ocampo E et al. Clinical, laboratory and imaging features of COVID-19: A systematic review and meta-analysis. Travel Med. Infect. Dis. 34, 1-13 (2020).

17. Feng X, Li S, Sun Q et al. Immune-Inflammatory Parameters in COVID-19 Cases: A Systematic Review and Meta-Analysis. Front. Med. 7, 1-14 (2020).

18. Tan L, Wang Q, Zhang D et al. Lymphopenia predicts disease severity of COVID-19: a descriptive and predictive study. Signal Transduct. Target Ther. 5(1), 1–3 (2020).

19. Henry BM, Aggarwal G, Wong J et al. Lactate dehydrogenase levels predict coronavirus disease 2019 (COVID-19) severity and mortality: A pooled analysis. Am J Emerg Med. In press (2020).

20. Sun Z, Zhang N, Li Y, Xu X. A systematic review of chest imaging findings in COVID-19. Quant. Imaging Med. Surg. 10(5), 1058–1079 (2020).

